# Identification of therapeutic targets for 18 clinical diseases by integrating human plasma proteome with genome

**DOI:** 10.1101/2023.10.23.23297376

**Authors:** Shifang Li, Meijiao Gong

**Author notes:** Correspondence: Shifang Li, Laboratory of Immunology and Vaccinology, FARAH, ULiège, Liège 4000, Belgium.

## Abstract

The proteome is an abundant source of potential therapeutic targets, and a comprehensive evaluation of proteins as therapeutic targets for a wide range of diseases is required. By screening 4,907 plasma proteins, we conducted a systematically two-sample proteome-wide Mendelian randomisation (MR) study to uncover potential therapeutic targets for 18 clinical diseases. Following MR analysis and stringent process filtering, a total of 146 causative plasma proteins (false discovery rate<0.05) were discovered. Colocalization analysis (Posterior Probability H4>0.8) further supported the causality of three proteins (MAP2K1, GFRA1, and THBS3) in gastroesophageal reflux disease; LYPLAL1 in keratoconus; three proteins (PCSK9, ANGPTL4, and GCKR) in familial hyperlipidemia; CRAT in atopic eczema; three proteins (IRF3, CA12, and TNFRSF1B) in hypothyroidism; AIF1 in age-related hearing impairment; SCARF2 in male pattern baldness; IRF3 in basal cell carcinoma; and four proteins (RPS6KA1, ULK3, MPPED2, and BTN3A1) in prostate cancer. Interestingly, having a genetically higher circulating CA12 level is associated with a lower risk of hypothyroidism (OR=0.47, *p*-value=1.68e-05, Posterior Probability H4=0.82). Single-cell RNA sequencing analysis showed that CA12 was mainly expressed in fibroblasts and epithelial cells in patient thyroid tissue and that its expression increased in older adults. Furthermore, with a proportion of 3.8%, hypothyroidism appears to mediate the effect of IRF3 on idiopathic pulmonary fibrosis, according to mediation analysis. Overall, our research could provide new insights into the etiology of clinical diseases as well as intriguing targets for the development of screening biomarkers and therapeutic treatments.

## Introduction

Plasma proteins are involved in numerous essential physiological processes, such as immunomodulation, substance transport, signaling, and homeostasis maintenance, and their dysregulation has been frequently reported in a variety of diseases, implying that these dysregulated proteins are involved in disease pathogenesis [1-2]. Given the importance of plasma proteins in disease and the fact that plasma proteins are a primary source of therapeutic targets, identifying disease-related plasma proteins can assist in deciphering disease pathophysiology and providing possible molecular targets for drug development [3-4]. Currently, large-scale proteomics study have uncovered over 18,000 protein quantitative trait loci (pQTLs) spanning over 4,800 proteins, including over 1,800 independent *cis*-pQTLs [5]. Through Mendelian randomisation (MR), these studies provide a significant data resource for systematically understanding the causal impact of plasma proteins on clinical disease. Normally, MR employs naturally randomized genetic diversity at the moment of conception as a natural experiment to uncover the causal relationship between exposure and disease, reducing the risk of reverse causation and confounding bias [6]. Proteome-wide MR has currently revealed important insights into the etiology of stroke, idiopathic pulmonary fibrosis (IPF), and COVID-19, as well as the prioritization of therapeutic targets [7-9]. Indeed, drugs containing human genetic evidence are more likely to succeed in Phase II and Phase III trials, with human genetic data supporting two-thirds of FDA-approved drugs in 2021 [10-12].

In this study, we utilized proteome-wide MR to identify circulating protein biomarkers linked with 18 clinical traits by integrating the human plasma proteome with genetic data. Colocalization analyses are conducted to determine the robustness of the proteins’ instrumental variables. Furthermore, we performed single-cell sequencing data analysis and mediation analysis to identify the enriched cell types of candidate proteins in tissues as well as the potential mechanisms mediated between diseases, respectively.

## Methods

### Proteomic data source

A large-scale protein quantitative trait loci (pQTL) study in 35,559 Icelanders yielded the largest and most thorough genome-wide association studies (GWAS) summary datasets on 4,907 circulating proteins [5]. The detailed description of the datasets can be found in the original study [5]. In brief, the plasma protein levels were determined using the SomaScan version 4 assay from SomaLogic, and the pQTL datasets were made up of associations between genome-wide genetic variations and plasma proteins that were adjusted for age and gender using the BOLT-LMM linear mixed model. As instrumental variables in the following MR analysis, genome-wide significant and independent single nucleotide polymorphisms (SNPs) with *p*-value<5e-08 and r^2^<0.001 of pQTLs were employed.

### 18 clinical traits data sources

To identify the potential therapeutic targets, the GWAS summary statistics from 18 diseases, including male pattern baldness (ID: GCST006661, 36,166 cases and 16,708 controls), gastroesophageal reflux disease (71,522 cases and 261,079 controls), keratoconus (ID: GCST90013442, 2,116 cases and 24,626 controls), selective IgA deficiency disease (ID: GCST003814, 1,635 cases and 4,852 controls), fibromuscular dysplasia (ID: GCST90026612, 1,556 cases and 7,100 controls), age-related hearing impairment (ID: GCST90012115, 125,688 cases and 205,071 controls), familial hyperlipidemia (ID: GCST90104003, 3,838 cases and 345,384 controls), atopic eczema (ID: GCST90027161, 22,474 cases and 774,187 controls), Brugada syndrome (ID: GCST90086158, 2,820 cases and 10,001 controls), cataract (ID: GCST009963, 21,679 cases and 387,283 controls), uterine fibroid (ID: GCST009158, 20,406 cases and 223,918 controls), amyotrophic lateral sclerosis (ID: GCST90027164, 27,205 cases and 110,881 controls), acne (ID: GCST90245818, 34,422 cases and 364,991 controls), posterior urethral valve (ID: GCST90134327, 756 cases and 4,823 controls), hypothyroidism (ID: GCST90204167, 51,194 cases and 443,383 controls), prostate cancer (ID: GCST90274714, 122,188 cases and 604,640 controls), basal cell carcinoma (ID: GCST90013410, 17,416 cases and 375,455 controls), and unipolar depression (ID: GCST006477, 66,809 individuals) were retrieved from the GWAS Catalog (https://www.ebi.ac.uk/gwas/) when an ID was provided (**Supplementary Table 1**). To the best of our knowledge, the exposure sample dataset (4,907 proteomic data) did not overlap with the outcome (18 clinical traits) we selected. Further information on the outcome dataset is listed in **Supplementary Table 1**. If standard error and effect size are missing but z-scores are included in the GWAS summary statistics file, effect size and standard error can be calculated as follows:

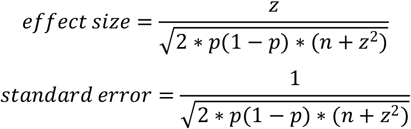

where n is the number of individuals used in the association study and p is the *p*-value for the association study.

### Mendelian randomisation analysis

Based on the presence of non-overlapping samples between exposure (4,907 proteome data) and outcome (18 clinical traits), two-sample MR analysis has been performed using the TwoSampleMR, as previously described [13-14]. In brief, the data were harmonized after clumping (r^2^<0.001) to exclude ambiguous SNPs with non-concordant alleles and palindromic SNPs. Following that, the Wald ratio method was used to generate effect estimates when considering a plasma protein instrumented by a single SNP, and the Inverse Variance Weighted (IVW) method was employed for proteins instrumented by two or more SNPs, followed by heterogeneity analysis. To test the consistency of the associations, additional methods such as simple mode, weighted mode, weighted median, and MR-Egger were applied. The MR-Egger intercept test was used to detect probable unbalanced pleiotropy (horizontal pleiotropy). The PhenoScanner (http://www.phenoscanner.medschl.cam.ac.uk/) databases were utilized to investigate whether variants had possible pleiotropic associations with other diseases or traits when fewer than three genetic instrumental factors were employed in MR analysis. Associations with *p*-value<5e-08 were deemed significant. In a heterogeneity test, we calculated I^2^ statistics with the “Isq()” function and heterogeneity *p*-value with the “mr_heterogeneity()” function; results with an I^2^> 50% and a heterogeneity *p*-value (Q_pval<0.05) were considered heterogeneous (substantial heterogeneity) [8]. The F-statistic was calculated to quantify instrument strength, and an F-statistic greater than 10 indicated an adequately powerful instrument. To account for multiple tests, the Benjamini-Hochberg correction, which governs the false discovery rate (FDR), was implemented. The association with a *p*-value of 0.05 but a Benjamini-Hochberg adjusted *p*-value greater than 0.05 was considered nominally significant, while the association with a Benjamini-Hochberg adjusted *p*-value greater than 0.05 was considered significant. Furthermore, the Steiger filtering approach was used to verify the validity of the causal direction between the hypothesized exposure and outcomes using the directionality_test() function in the “TwoSampleMR” package [14]. In addition, we performed a leave-one-out analysis to examine whether the MR estimates could be explained by a single SNP (removing estimates where all but one leave-one-out configuration had a *p*-value<0.05). Only MR results that met all of the following criteria were chosen to avoid complex causation relationships: (1) MR-Egger regression revealed no pleiotropy (*p*-value>0.05); (2) true causal direction and Steiger *p*-value*<*0.05; (3) heterogeneity test I^2^<0.5; (4) leave-one-out analysis MR *p*-value*<*0.05 after removing outliers; and (5) FDR*<*0.05.

### Colocalization Analysis

Colocalization analysis was executed to figure out whether the protein and diseases shared the same causative genetic variations and to rule out confounding owing to linkage disequilibrium (LD). This was performed using the “coloc” R package [15]. The following five mutually exclusive hypotheses were tested: (1) Neither protein nor disease has a causal SNP (H0); (2) only protein has a causal SNP (H1); (3) only disease has a causal SNP (H2); (4) both protein and disease have a causal SNP, but the two causal SNPs differ (H3); and (5) both protein and disease have a causal SNP and share the same SNP (H4). To measure the support for each hypothesis, the posterior probability was adopted. A posterior probability for H4 (PH4) of more than 80% was regarded as significant evidence of colocalization.

### Single-cell RNA sequencing analysis

Single-cell RNA-sequencing of 54,762 cells taken from normal thyroid tissue from 7 individuals who had thyroidectomy and had verified instances of differentiated thyroid carcinoma were retrieved from Gene Expression Omnibus (https://www.ncbi.nlm.nih.gov/geo/, GSE182416) [16]. Seurat 4.2.0 was used for processing and analyzing the single-cell dataset, as previously stated [13]. In short, low-quality cells by selecting feature numbers ranging from 200 to 3,000 were excluded. The data were normalized using the “LogNormalize” method using the “NormalizeData” function. The “vst” selection method in the “FindVariableFeatures” function yielded a total of 2,000 highly variable genes for each sample. The “FindIntegrationAnchors” function was utilized to correct the batch effect and merged the 7 patients for further analysis. The k-nearest neighbors were determined using principal component analysis (PCA). For data visualization, the dimension reduction approach UMAP (Uniform Manifold Approximation and Projection) was utilized. Cell type annotation was carried out in their study based on the specific gene expression [16].

### Mediation analysis

The product of coefficients approach was employed to quantify the impacts of proteins on IPF outcomes via hypothyroidism for proteins that causally are associated with IPF and hypothyroidism, as mentioned by Yoshiji and her colleagues [8]. In brief, the effect of IRF3 on hyperthyroidism was first evaluated and then multiplied by the effect of hyperthyroidism on IPF. Following that, the proportion of the entire effect of IRF3 on IPF mediated by hyperthyroidism was calculated by dividing the hyperthyroidism-mediated effect (hyperthyroidism-to-IPF) by the overall effect (IRF3-to-IPF) [8,17].

## Results and discussion

### Identification of causal proteins associated with clinical traits

The MR analysis was used to evaluate the causal effects of 4,907 plasma proteins on 18 diseases by using *cis*-pQTLs (pQTLs that reside within a 1 Mb region around a transcription start site of a protein-coding gene) for these proteins as instrumental variables and 18 clinical traits as outcomes (**Figure 1A** and **Supplemental Table 1**). Given that *cis*-pQTLs are more likely than *trans*-pQTLs to directly alter the transcription or translation of their linked gene, the likelihood of directional horizontal pleiotropy is considerably reduced [8]. Following MR analysis and stringent process filtering (**Method** and **Figure 1A**), a total of 146 plasma proteins were found for associations with 18 clinical traits (**Figure 1B** and **Supplemental Table 1**). Particularly, the causal associations were found for 12 clinical traits, including male pattern baldness (6 proteins), gastroesophageal reflux disease (15 proteins), Keratoconus (3 proteins), selective IgA deficiency disease (2 proteins), age-related hearing impairment (12 proteins), familial hyperlipidemia (8 proteins), atopic eczema (4 proteins), Brugada syndrome (1 protein), amyotrophic lateral sclerosis (1 protein), hypothyroidism (27 proteins), basal cell carcinoma (5 proteins), and prostate cancer (62 proteins). For example, a genetic tendency to elevated LYPLAL1 was linked to an increased risk of keratoconus (OR per-1-SD higher plasma protein level [95% confidence interval (CI)]=2.52[1.56, 4.06]; *p*-value*=*0.00014). Higher levels of genetically predicted CRAT were linked to an increased incidence of atopic eczema (OR[95% CI]=1.63[1.27, 2.10]; *p*-value=0.00012). Genetically determined higher SCARF2 levels were associated with a higher risk of male pattern baldness (OR[95%CI]: 1.23[1.11, 1.36]; *p*-value=3.91e-05). It is worth noting that the causative protein that is significantly linked to one clinical trait may also be linked to other clinical traits, while this is nominally significant (**Figure 1B**). For example, THBS3, which was associated with an increased risk of gastroesophageal reflux disease (OR[95%CI]=1.31[1.15, 1.49]; *p*-value=6.2e-05), was also linked to a higher risk of cataract (OR[95%CI]=1.014[1.003, 1.025]; *p*-value=0.0125) and uterine fibroid (OR[95%CI]=1.32[1.05, 1.67]; *p*-value=0.016). Furthermore, CRAT, which was linked to atopic eczema, was also associated with a higher risk of age-related hearing impairment (OR[95%CI]=1.079[1.022, 1.139]; *p*-value=0.0059) and fibromuscular dysplasia (OR[95%CI]=3.28[1.11,9.71]; *p*-value=0.031). These findings indicated an opportunity for existing pharmaceutical repurposing in the prevention of disease. In fact, inhibitors of interleukin-23 (IL-23) signaling, initially developed for psoriasis, have been repurposed for Crohn’s disease based on GWAS that found links between genetic variants in the IL23R gene and Crohn’s disease [3,18,19]. Similarly, IL-17A signaling inhibitors established for psoriasis, rheumatoid arthritis, and uveitis have been investigated and approved for ankylosing spondylitis based on GWAS findings [3]. Notably, LYPLAL1, which has been connected to an increased risk of keratoconus, has also been linked to a lower risk of male pattern baldness (OR[95%CI]=0.87[0.77, 0.995]; *p*-value=0.043) and atopic eczema (OR[95%CI]=0.74[0.60, 0.92]; *p*-value=0.0062). These findings suggested that when repurposing drugs, it is essential to consider the potential effects caused by multiplicity.

**Figure 1.**
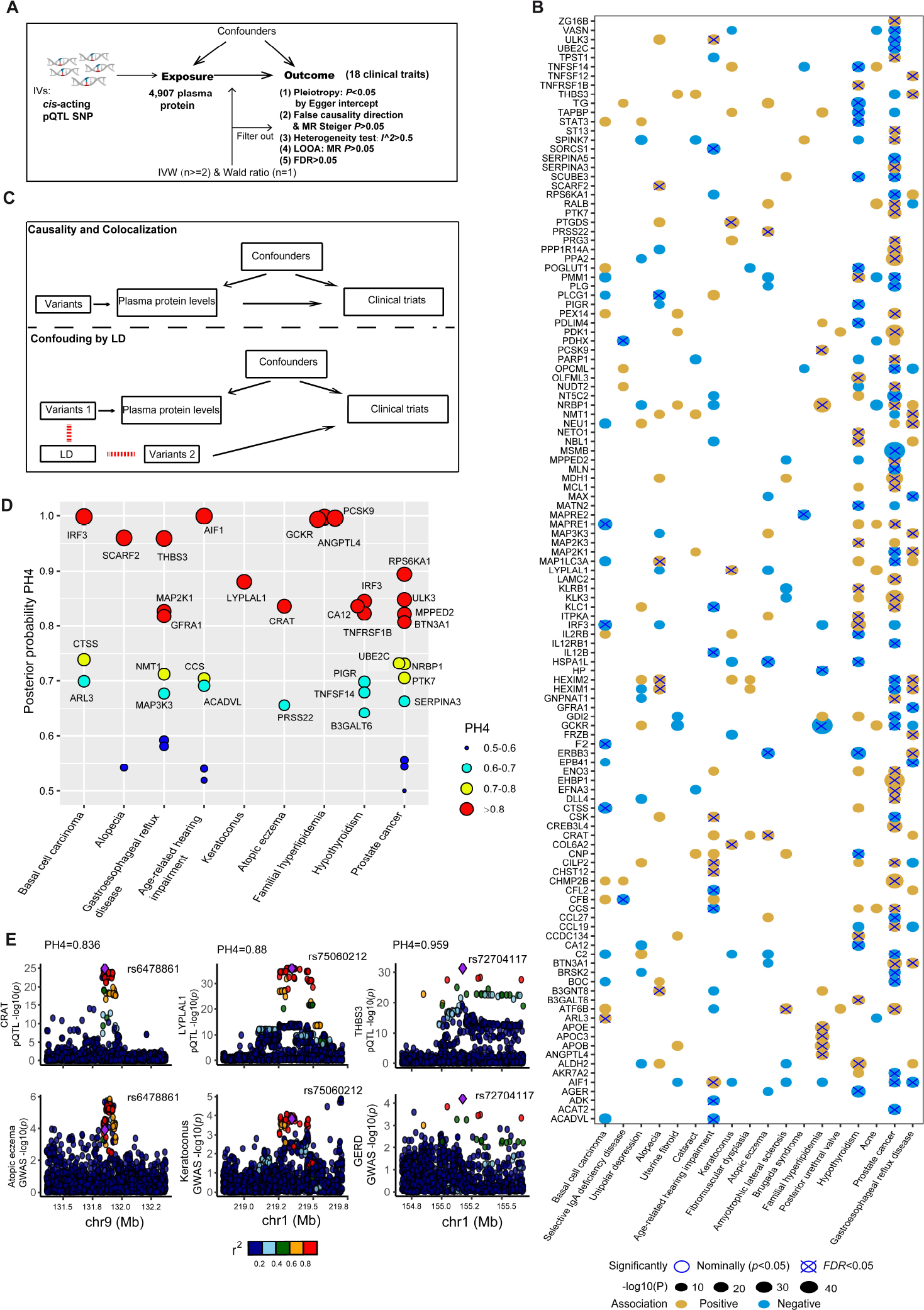
Mendelian randomisation and colocalization results. (**A**) A diagram depicting the Mendelian randomization principle and procedure filtering for MR results. LOOA, leave-one-out analysis. (**B**) Shared the associations between 146 causative plasma proteins (related with at least one clinical trait) and 18 clinical traits from MR analysis. For visualizing, only MR results showing no pleiotropy (MR-Egger, *p*-value>0.5), no heterogeneity (I^2^<0.5), and true causality direction (Steiger test, *p*-value<0.05) were included. (**C**) The two mathematical models of genetic colocalization, causation, and colocalization, as well as LD confounding. (**D**) Scanning for colocalization evidence of causative plasma proteins with outcomes. The size of the circle represents the posterior probability for H4, and the color of the circle represents the classification of the evidence. Red indicates high evidence for colocalization (PH4>0.8); yellow indicates medium evidence for colocalization (PH4>0.7); and green and black indicate moderate evidence for colocalization (PH4<0.7). (**E**) The locus-compare scatter plot for the CRAT, LYPLAL1, and THBS3 association signals. The findings of colocalization analyses for CRAT (left), LYPLAL1 (center), and THBS3 (right) are shown. The marked SNP represents the genetic variants used for the MR analysis.

Colocalization analysis was then implemented to determine whether previously identified relationships between proteins and clinical traits were driven by linkage disequilibrium (LD) (**Figure 1C**). Among the 146 causative proteins, 18 showed higher colocalization evidence (posterior probability PH4>0.8), whereas 6 showed medium colocalization evidence (posterior probability PH4>0.7) (**Figure 1D**). For example, the colocalization indicated that the genetic variations linked to CRAT, LYPLAL1, and THBS3 (*cis*-pQTLs) were caused by the same genetic variants that underpin the association with individual clinical traits (**Figure 1E**). For the other 122 causative proteins, there was no colocalization evidence (posterior probability PH4<0.6), indicating that the MR findings for these proteins were likely biased by LD.

### Genetically increased circulating CA12 level is associated with a reduced risk of hypothyroidism

Based on the MR and colocalization analysis results, the causative protein related to the clinical traits was sought to further examine the role they played. In the case of hypothyroidism, among the three causative proteins, a one standard deviation rise in genetically predicted CA12 levels was linked with a lower risk of hypothyroidism (OR=0.47, 95%CI 0.33-0.66, *p*-value=1.68e-05) (**Figure 1B, Figure 2A** and **Supplementary Tables 1**). To confirm the premise of a lack of directional pleiotropy, which can reintroduce confounding, we used PhenoScanner (http://www.phenoscanner.medschl.cam.ac.uk/) to find out whether the CA12 *cis*-pQTLs were related to any characteristics or diseases at the genome-wide significant threshold of *p*-value<5e-08. The deCODE study’s primary *cis*-pQTL for CA12 (rs7183733) was linked to dentures (**Supplementary Tables 1**). Indeed, employing CA12 *cis*-pQTL as exposure and dentures as outcomes, MR analysis revealed that CA12 levels were expected to impact dentures (OR=1.12, 95%CI 1.08-1.16, *p*-value=5.59e-12) with no reverse causation (*p*-value=1.83e-04, MR-Steiger test). This finding implies that the association between CA12 and dentures is a case of vertical pleiotropy, which does not contradict MR explanation. GTEx v8 (https://gtexportal.org/), which contains expression data from 49 tissues and 838 individuals, was used to investigate the tissues in which CA12 is expressed. When compared to whole blood, CA12 was highly expressed in numerous tissues, including the thyroid (*p*-value<0.001) (**Figure 2B**). To better understand the cell type of origin of CA12, we examined single-cell CA12 expression in human thyroid tissues from thyroidectomy patients (GSE182416, available at https://www.ncbi.nlm.nih.gov/geo/) [16]. CA12 was highly enriched in thyroid epithelial cells and fibroblasts in single-cell sequencing as compared to other cell types in adipose tissues (*p*-value<0.001) (**Figure 2C**). No difference in expression between epithelial cells and fibroblasts was found (*p*-value>0.05). These findings suggested that certain cell types may be accountable for local CA12 production in these tissues. Furthermore, we noticed that CA12 was considerably more expressed in older persons (*p*-value<0.001) (**Figure 2D**), who had a greater prevalence and incidence of hypothyroidism than young ones [20]. Taken together, these findings imply that CA12 may be an appropriate target for hypothyroidism.

**Figure 2.**
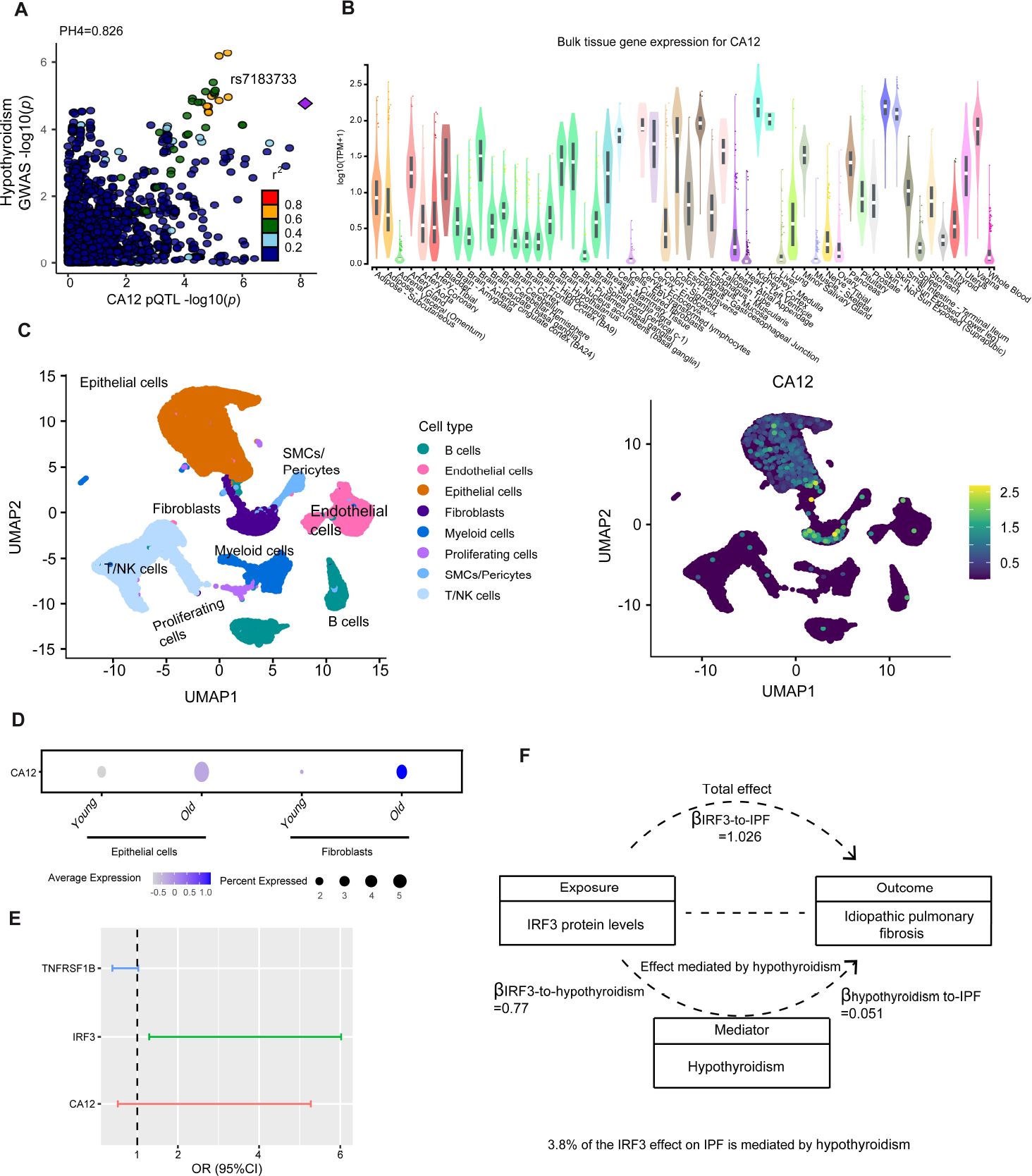
Follow-up analyses for causal proteins associated with hypothyroidism. (**A**) The locus-compare scatter plot compares the quantitative trait loci (pQTL) of CA12 and the GWAS of hypothyroidism. (**B**) CA12 expression profile in human tissues in GTEx v8 (https://gtexportal.org/). CA12 expression levels were displayed on a log transcript per thousand plus one (TPM+1) scale. (**C**) CA12 expression patterns in human thyroid tissues from thyroidectomy patients (GSE182416, accessible at https://www.ncbi.nlm.nih.gov/geo/). NK cells: natural killer cells; SMCs: smooth muscle cells. (**D**) A dot-plot revealing the expression of CA12 in epithelial cells and fibroblasts in children and adults. (**E**) Forest plots for the effect of three selected proteins on IPF. (**F**) Mediation effects of IRF3 on IPF via hypothyroidism. For the effect of IRF3 on IPF mediated by hypothyroidism, the product of coefficients method calculates the proportion mediated by multiplying β_IRF3-to-hypothyroidism_ and β_hypothyroidism-to-IPF_ and subsequently dividing it by β_IRF3-to-IPF_, where β_IRF3-to-hypothyroidism_ is the effect of IRF3 on hypothyroidism, β_hypothyroidism-to-IPF_ is the effect of hypothyroidism on IPF, and β_IRF3-to-IPF_ is the total effect of IRF3 on IPF.

In fact, the causative protein found in the study might be utilized to infer disease mediator associations [7]. As an example, recent research has shown that hypothyroidism has a positive causal effect on IPF [21]; however, the mechanisms underlying this association are not well understood. To evaluate the indirect effect of proteins on IPF outcomes via hypothyroidism, we performed a mediation analysis utilizing the effect estimates from two-step MR and the overall effect from primary MR. To accomplish this, the largest GWAS of IPF (2,668 cases and 8,591 controls) was extracted from a recent study [22] and three proteins (IRF3, CA12, and TNFRSF1B) that have a causal effect on hypothyroidism were initially used to identify a causal effect on IPF using MR analysis, as described above. Surprisingly, only IRF3 was found to have a causal effect on IPF (*p*-value<0.05/3=0.016, Bonferroni-adjusted for three proteins) (OR=2.79, 95%CI 1.29-6.02, *p*-value=0.0088) (**Figure 2E** and **Supplementary Tables 1**). Following that, the product of coefficients method was utilized to assess the mediation effect, and 3.8% of the IRF3 mediation affect on IPF via hypothyroidism was uncovered (**Figure 2F**).

Overall, our analysis identified 18 putative causative proteins for clinical traits by integrating proteogenomics research. More research is needed to determine the viability of the 18 discovered proteins as therapeutic targets for clinical disease therapy. Furthermore, the detailed mechanism by which these proteins alter clinical traits should be explored in depth using a multi-omics approach.

## Supporting information

Supplementary Table 1 The summary statistics obtained in this study.

## Data Availability

All the data produced in the present work is contained in the manuscript

## Competing interests

None declared.

## Author contributions

Conception and design: SF. Data analyses: SF. Manuscript writing: SF. Data acquisition: SF and MJ.

## Acknowledgments

We would like to thank the IPF GWAS Collaborative Group for providing us with the IPF GWAS summary data. The authors would also like to thank the other researchers who contributed to developing the GWAS datasets utilized in this work for making them available for research purposes.

## Notes

### Competing Interest Statement

The authors have declared no competing interest.

### Funding Statement

This study did not receive any funding

### Summary of Updates

The results of proteome-wide Mendelian randomisation and colocalization analyses for prostate cancer and basal cell carcinoma have been added to the manuscript.

